# Efficacy and safety of a third SARS-CoV-2 vaccination in multiple sclerosis vaccine non-responders

**DOI:** 10.1101/2021.10.15.21264977

**Authors:** Marton König, Hilde Marie Torgauten, Mathias Herstad Øverås, Adity Chopra, Åslaug Rudjord Lorentzen, The Trung Tran, Siri Mjaaland, Ingeborg Sundsvalen Aaberge, Kjell-Morten Myhr, Stig Wergeland, Tone Berge, Hanne Flinstad Harbo, Øivind Fredvik Torkildsen, Trygve Holmøy, Elisabeth Gulowsen Celius, Ludvig Andre Munthe, John Torgils Vaage, Fridtjof Lund-Johansen, Gro Owren Nygaard

## Abstract

**Importance:** Vaccination against severe acute respiratory syndrome coronavirus 2 (SARS-CoV-2) to protect against coronavirus disease of 2019 (COVID-19) is recommended for patients with multiple sclerosis (pwMS). However, approximately 80% of all pwMS treated with anti-CD20 therapy (rituximab, ocrelizumab) or fingolimod have low or absent humoral immunity after vaccination with two doses of SARS-CoV-2 mRNA vaccines. The efficacy and safety of a third vaccine dose in this group is largely unknown.

**Objective:** To characterize the humoral immunogenicity and the safety of a third dose of mRNA-COVID-19 vaccine in anti-CD20-or fingolimod-treated pwMS with low or absent humoral immunity (i.e., anti-SARS-CoV-2 IgG <70 arbitrary units (AU) and <5 AU, respectively) after two vaccinations.

**Design, setting and participants:** 130 anti-CD20- or fingolimod-treated pwMS with low or absent humoral immunity despite full vaccination against SARS-CoV-2, received a third dose of SARS-CoV-2 mRNA vaccine. Humoral immunity (i.e., antibody response against SARS-CoV-2) and the frequency and characteristics of side-effects were analyzed in all participants.

**Exposures:** A third vaccine dose against SARS-CoV-2 with BNT162b2- or mRNA-1273-COVID-19 vaccine.

**Main outcomes and measures:** Patient- and treatment-specific variables were acquired using a digital questionnaire, the Norwegian Immunization Registry and hospital journals. Humoral immunity was assessed by measuring SARS-CoV-2 SPIKE receptor-binding domain (RBD) IgG response. Low/absent humoral immunity was assumed in cases of AU<70 after anti-SPIKE protein-based serology 3-5 weeks after revaccination.

**Results:** A third dose of SARS-CoV-2 mRNA vaccine increased anti-SARS-CoV-2 SPIKE RBD IgG levels significantly. The proportion of patients with assumed protective humoral immunity (anti-SARS-CoV-2 SPIKE RBD IgG > 70 AU) were 25% among patients using anti-CD20 therapy and 7% among those treated with fingolimod. No adverse events were registered during the study period.

**Conclusion and relevance:** A third dose of mRNA-COVID-19 vaccine was associated with significantly increased levels of anti-SARS-CoV-2 SPIKE RBD IgG, – and hence assumed protective humoral immunity - in anti-CD20- or fingolimod-treated pwMS with low or absent humoral immunity despite full vaccination. The effect of a third vaccine dose was limited and more prominent among those treated with anti-CD20 therapy.

## Introduction

Severe acute respiratory syndrome coronavirus 2 (SARS-CoV-2) corona disease of 2019 (COVID-19) is associated with high mortality and detrimental effects on society, economy, and individual lives. Patients with multiple sclerosis (pwMS) do not have an increased risk of SARS-CoV-2 infection or severe COVID-19 disease per se, however, the risk is elevated in the presence of comorbidities, higher age, greater MS-associated disability, progressive disease course, and ongoing treatment with certain disease-modifying therapies (DMT) (1-4). Early initiation of treatment with high-efficacy DMT is the single most important factor in reducing long-term disability in pwMS (5). Specific DMTs are, however, associated with an increased risk of infections (6). Expert organizations worldwide recommend that all pwMS should be vaccinated against COVID-19 (7). There is increasing evidence of reduced humoral immunity after two doses of mRNA-COVID-19 vaccines among patients treated with anti-CD20 therapy (rituximab, ocrelizumab) or fingolimod (8, 9). A beneficial effect of three doses of mRNA-COVID-19 vaccines has been observed in solid-organ transplant recipients (10). However, the effect of a third vaccine dose on anti-SARS-Cov 2 IgG antibody levels in pwMS is largely unknown. We report results from a study designed to assess the immunogenicity and the safety of a third dose of COVID-19 mRNA vaccine in pwMS treated with anti-CD20 therapy or fingolimod.

## Methods

The study was approved by the Norwegian, South-Eastern Regional Ethical Committee (Ref. Nr. 200631) and the Norwegian Medicines Agency (EudraCT Number: 2021-003618-37). All participants gave written, informed consent.

### Study population

All anti-CD20- or fingolimod-treated pwMS with low or absent humoral immunity after two mRNA-COVID-19 vaccine doses were invited to the study. Inclusion criteria were MS diagnosis, signed informed consent, completed COVID-19-vaccination (i.e., two vaccine doses or past COVID-19 and one vaccine dose) and SARS-CoV-2 SPIKE RBD IgG <70 arbitrary units (AU) 3-12 weeks after full vaccination.

### Testing of humoral immune response

Antibodies to full length Spike (HexaPro) from SARS-CoV-2 and the receptor-binding domain (RBD) were measured using a bead-based flow cytometric assay (11) in all included patients 3-12 weeks after full vaccination and 3-5 weeks after revaccination. Post-immunization IgG titers were used as a correlate of protection (12), and reduced immunity was assumed in cases of IgG < 70 AUcorresponding to a lower level than found in 99% of healthy vaccinated subjects (9). IgG levels < 5 AU were defined as no antibody response, while IgG levels between 5-70 AU were defined as weak antibody response. Calibration to the WHO international standard showed that 70 AU corresponds to approximately 40 Binding Antibody Units per milliliter (BAU/ml).

### Data collection

Demographic, disease-, treatment- and vaccination-specific variables were acquired through a digital questionnaire completed by all patients and from patient journals. Information regarding side-effects were collected 3-5 weeks after revaccination. Information regarding COVID-19-vaccines was extracted from the Norwegian Immunization Registry.

### Statistics

Continuous and categorical variables were compared using Mann-Whitney and Fisher’s exact tests, respectively. A *p* value less than 0.05 was considered statistically significant. Correlations were assessed by Spearman p. Statistical analysis was conducted using SPSS^®^ version 26 (SPSS Inc., Chicago, USA).

## Results

Of 175 invited pwMS, 130 patients (100 using rituximab, 1 ocrelizumab, and 29 fingolimod) had signed informed consent, completed COVID-19-revaccination (with a third dose of mRNA-COVID-19 vaccine), filled out digital questionnaire regarding side-effects, and underwent testing of humoral immune response before October 1^st^ 2021. Demographic and background variables are described in Table 1.

**Table 1.** Clinical and demographic variables of patients with multiple sclerosis who underwent revaccination.

| Study population | All patients<br>n = 130 | anti-CD20<br>n = 101 | Fingolimod<br>n = 29 |
| --- | --- | --- | --- |
| Gender, n (%) |  |  |  |
| Females | 97 (75) | 80 (79) | 17 (59) |
| Males | 33 (25) | 21 (21) | 12 (41) |
| Age, years |  |  |  |
| Median | 47.5 | 47.2 | 48.9 |
| 25-75 IQR | 40.6-56 | 40-60.8 | 41.8-56.9 |
| Disease duration, years |  |  |  |
| Median | 6.7 | 5.1 | 14.1 |
| 25-75 IQR | 2.6-13.6 | 2.6-11.6 | 7.6-18.6 |
| Disability by EDSS |  |  |  |
| Median | 2.0 | 2.0 | 2.0 |
| 25-75 IQR | 1-3.1 | 1-3.1 | 1-3.3 |
| Multiple sclerosis type, n (%) |  |  |  |
| Relapsing remitting | 114 (88) | 85 (84) | 29 (100) |
| Progressive | 17 (12) | 16 (16) | 0 |
| SARS-CoV-2 RBD IgG titer after full-vaccination (AU) |  |  |  |
| Median (Mean) | 3 (9) | 3 (8.9) | 5 (9.2) |
| 25-75 IQR | 2-28.8 | 2-7 | 2.6-8.5 |
| < 5 AU, n (%) | 80 (62) | 67 (66) | 13 (45) |
| 5-70 AU, n (%) | 50 (38) | 34 (34) | 16 (55) |
| > 70 AU, n (%) | 0 (0) | 0 (0) | 0 (0) |
| SARS-CoV-2 RBD IgG titer after revaccination |  |  |  |
| Median (Mean) | 6 (44) | 4 (49.4) | 13 (25.1) |
| 25-75 IQR | 3-59.3 | 2-69.5 | 4-38 |
| < 5 AU, n (%) | 59 (45) | 51 (50) | 8 (28) |
| 5-70 AU, n (%) | 44 (34) | 25 (25) | 19 (66) |
| > 70 AU, n (%) | 27 (21) | 25 (25) | 2 (7) |
| Time from last Tx dose to revaccination, months |  |  |  |
| Median |  | 4 |  |
| 25-75 IQR |  | 2.1-5.6 |  |
| Absolute lymphocyte count (cells/mm<sup>3</sup>) |  |  |  |
| Median | 1300 | 1500 | 550 |
| 25-75 IQR | 900-1700 | 1200-1800 | 425-775 |
| CD19-B-cell count (cells/mm<sup>3</sup>) |  |  |  |
| Median |  | 0 |  |
| 25-75 IQR |  | 0-2.3 |  |
| Tx, treatment; IQR, Interquartile range |  |  |  |

While the majority of pwMS received the BNT162b2 vaccine (83% as the first, and 85% as the second jab) upon full vaccination, 85% received revaccination with the mRNA-1273 vaccine. The mean time between the last jab of full vaccination and revaccination was 94 days (STD 30.8 days) and 78 days (STD 18.2 days) in patients treated with anti-CD20 and fingolimod, respectively.

Mean levels of anti-SARS-CoV-2 SPIKE RBD IgG increased significantly in both treatment groups (anti-CD20: p < 0.001; fingolimod: p = 0.006) after revaccination. The proportion of patients with assumed protective humoral immunity (anti-SARS-CoV-2 SPIKE RBD IgG > 70) were 25% among patients using anti-CD20 therapy and 7% among those treated with fingolimod after revaccination (Figure 1). Higher absolute lymphocyte (regardless of treatment) and CD19-B-cell counts (in patients receiving anti-CD20 therapy) were significantly associated with the development of protective humoral immunity (p = 0.03 in both cases).

**Figure 1.**
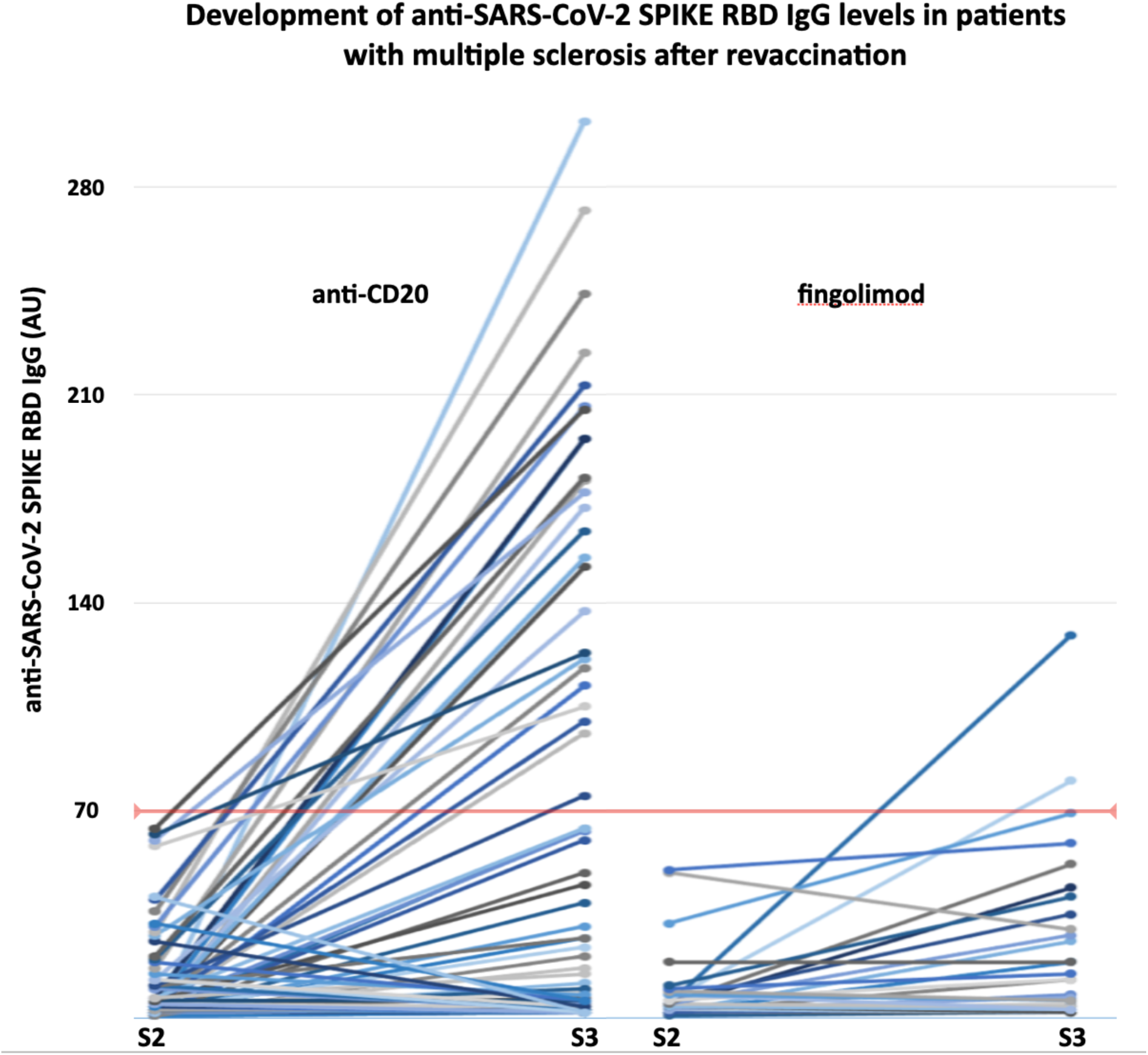
The development of anti-SARS-CoV-2 SPIKE RBD IgG levels in anti-CD20 or fingolimod treated patients with multiple sclerosis undergoing revaccination. S2-3, antibody sample after second and third vaccine dose, respectively.

Side-effects were observed in 63% of pwMS treated with anti-CD20 therapy and in 38% treated with fingolimod. The most common side-effects were transient local pain and fatigue in both groups. No patients experienced serious adverse effects after revaccination (Table 2). The mean absolute lymphocyte count was significantly higher in patients reporting side-effects, compared to those who did not (p = 0.03).

**Table 2.** Vaccination specific variables of patients with multiple sclerosis who underwent COVID-19 revaccination.

| Study population | All patients<br>n = 130 | anti-CD20<br>n = 101 | Fingolimod<br>n = 29 |
| --- | --- | --- | --- |
| Type of V1, n (%) |  |  |  |
| BNT162b2 | 107 (83) | 87 (86) | 20 (69) |
| mRNA-1273 | 18 (14) | 10 (10) | 8 (28) |
| ChAdOx1-S | 4 (3) | 3 (3) | 1 (3) |
| Type of V2, n (%) |  |  |  |
| BNT162b2 | 111 (85) | 90 (89) | 21 (72) |
| mRNA-1273 | 19 (15) | 11 (11) | 8 (28) |
| Time between V2 and S2 (days) |  |  |  |
| Median | 39 | 53 | 28 |
| 25-75 IQR | 23-77 | 25-81.5 | 21-36.8 |
| Time between V2 and V3 (days) |  |  |  |
| Median | 84 | 91 | 77 |
| 25-75 IQR | 71.5-109 | 73.5-113.5 | 67.5-83 |
| Type of V3, n (%) |  |  |  |
| BNT162b2 | 19 (15) | 12 (12) | 7 (24) |
| mRNA-1273 | 111 (85) | 89 (88) | 22 (76) |
| Time between V3 and S3 (days) |  |  |  |
| Median | 22 | 22 | 23 |
| 25-75 IQR | 21-25 | 21-24.5 | 21-35 |
| Side effects, n (%) |  |  |  |
| Total | 75 (58) | 64 (63) | 11 (38) |
| Local pain | 61 (47) | 54 (54) | 7 (24) |
| Local itch | 8 (6) | 8 (8) | 0 (0) |
| Local rubor | 15 (12) | 15 (15) | 0 (0) |
| Fatigue | 52 (40) | 44 (44) | 8 (28) |
| Headache | 42 (32) | 39 (39) | 3 (10) |
| Muscle pain | 47 (36) | 41 (41) | 6 (21) |
| Nausea | 11 (9) | 9 (9) | 2 (7) |
| Airway symptoms | 10 (8) | 9 (9) | 1 (3) |
| Feeling of sickness | 33 (25) | 33 (33) | 0 (0) |
| Insomnia | 14 (11) | 13 (13) | 1 (3) |
| Fever | 27 (21) | 25 (25) | 2 (7) |
| Lymphedema | 5 (4) | 4 (4) | 1 (3) |
| Seeked medical help | 4 (3) | 4 (4) | 0 (0) |
| Hospitalisation | 0 (0) | 0 (0) | 0 (0) |
| MS-relapse | 0 (0) | 0 (0) | 0 (0) |
| IQR, Interquartile range; V1/2/3, first/second/third vaccine dose;<br>S2-3, antibody sample after V2 and V3, respectively |  |  |  |

## Discussion

We present the first results of immune responses to COVID-19 revaccination in pwMS treated with anti-CD20 therapy or fingolimod. Our results show that a third dose of mRNA-COVID-19 vaccine was associated with modestly increased levels of anti-SARS-CoV-2 SPIKE RBD IgG antibodies in patients with reduced protective humoral immunity prior to reimmunization. Revaccination increased the proportion of patients with protective humoral immunity in both groups, and this effect was more pronounced among patients using anti-CD20 therapy.

Earle and colleagues found a robust correlation between neutralizing IgG titer and efficacy and between binding antibody titer and efficacy, despite geographically diverse study populations, and despite use of different endpoints, assays, convalescent sera panels and manufacturing platforms (12). Together with evidence from natural history studies and animal models, their results support the use of post-immunization IgG titers as the basis for establishing a correlate of protection for COVID-19 vaccines.

Only 21% of our patients developed antibody levels where protective immunity can be assumed, hence, the clinical significance and the effect of a third vaccine dose remains unclear. On the other hand, it may be important that 25% of those treated with anti-CD20 therapy acquired assumed protective humoral immunity, as these patients have an approximately three-fold risk of developing serious COVID-19 (2).

Patients with higher absolute lymphocyte counts seems to have a better response and pwMS with anti-CD20 therapies should probably be revaccinated shortly before the next treatment course is planned (9). If disease activity allows, one might also consider widening treatment intervals to increase the probability of response to vaccination.

The rate of side-effects was higher in pwMS using anti-CD20 therapy compared to those treated with fingolimod, however, no serious adverse events were encountered and revaccination can be considered safe.

## Limitations

Our results are based on short-term follow-up of a limited number of patients. Long-term follow-up is undoubtedly of great importance, especially when the endpoints (e.g., antibody titers) are dynamic. Furthermore, we only report data regarding IgG responses as a correlate of humoral immunity, while the adaptive immune response to SARS-CoV-2 seems to depend also on cellular responses (13). Finally, we do not report on neither the durability of the response or the clinical effect of revaccinations.

## Conclusions

Revaccination (i.e., administration of a third dose of mRNA-COVID-19 vaccine) of anti-CD20- or fingolimod-treated pwMS with low antibody levels after two mRNA-COVID-19 vaccine doses improves protective humoral immunity. The effect of a third vaccine dose is limited and more prominent among those treated with anti-CD20 therapy. Our results show that revaccination can be considered safe, and hence, indicated to reduce the risk of serious COVID-19.

## Data Availability

All data produced in the present study are available upon reasonable request to the authors
All data produced in the present work are contained in the manuscript.

## Acknowledgements

We express our gratitude to Lars Skattebøl, Einar August Høgestøl, Rebecca Cox, the Bergen COVID-19 Group, and the Coalition for Epidemic Preparedness Innovations for their cooperation and support.

## Disclosures

M. König has received speaker honoraria from Novartis, Biogen and AstraZeneca.

H.M. Torgauten has nothing to disclose.

M.H. Øverås has nothing to disclose.

A. Chopra has nothing to disclose.

Å. Lorentzen research fundings from Sanofi.

T. Tran has nothing to disclose.

S. Mjaaland has nothing to disclose.

I.S. Aaberge has nothing to disclose.

K.M. Myhr has received unrestricted research grants to his institution; scientific advisory board and speaker honoraria from Biogen, Merck, Novartis, Roche and Sanofi, and has participated in clinical trials organized by Biogen, Merck, Novartis, Roche and Sanofi S. Wergeland has received speaker honoraria from and served on scientific advisory boards for Biogen, Janssen-Cilag, Sanofi and Novartis.

T. Berge has received unrestricted research grants from Biogen Idec and Sanofi Genzyme.

H.F. Harbo has received honoraria for lecturing or advice from Biogen, Merck, Roche, Novartis and Sanofi.

Ø.F. Torkildsen has received speaker honoraria from and served on scientific advisory boards for Biogen, BMS, Jansen, Sanofi, Merck and Novartis.

T. Holmøy has received speaker honoraria and/or unrestricted research grants from Biogen, Merck, Roche, Novartis, Sanofi and Bristol-Myers Squibb, and participated in clinical trials organized by Merck, Sanofi and Roche.

E.G. Celius has received honoraria for lecturing and advice from Biogen, BMS, Janssen, Merck, Novartis, Roche and Sanofi.

L.A. Munthe has nothing to disclose.

J.T. Vaage has nothing to disclose.

F. Lund-Johansen has nothing to disclose.

G.O. Nygaard has nothing to disclose.

